# A Machine Learning Study of 534,023 Medicare Beneficiaries with COVID-19: Implications for Personalized Risk Prediction

**DOI:** 10.1101/2020.10.27.20220970

**Authors:** Chen Dun, Christi M. Walsh, Sunjae Bae, Amesh Adalja, Eric Toner, Timothy A. Lash, Farah Hashim, Joseph Paturzo, Dorry L. Segev, Martin A. Makary

## Abstract

**Background:** Global demand for a COVID-19 vaccine will exceed the initial limited supply. Identifying individuals at highest risk of COVID-19 death may help allocation prioritization efforts. Personalized risk prediction that uses a broad range of comorbidities requires a cohort size larger than that reported in prior studies.

**Methods:** Medicare claims data was used to identify patients age 65 years or older with diagnosis of COVID-19 between April 1, 2020 and August 31, 2020. Demographic characteristics, chronic medical conditions, and other patient risk factors that existed before the advent of COVID-19 were identified. A random forest model was used to empirically explore factors associated with COVID-19 death. The independent impact of factors identified were quantified using multivariate logistic regression with random effects.

**Results:** We identified 534,023 COVID-19 patients of whom 38,066 had an inpatient death. Demographic characteristics associated with COVID-19 death included advanced age (85 years or older: aOR: 2.07; 95% CI, 1.99-2.16), male sex (aOR, 1.88; 95% CI, 1.82-1.94), and non-white race (Hispanic: aOR, 1.74; 95% CI, 1.66-1.83). Leading comorbidities associated with COVID-19 mortality included sickle cell disease (aOR, 1.73; 95% CI, 1.21-2.47), chronic kidney disease (aOR, 1.32; 95% CI, 1.29-1.36), leukemias and lymphomas (aOR, 1.22; 95% CI, 1.14-1.30), heart failure (aOR, 1.19; 95% CI, 1.16-1.22), and diabetes (aOR, 1.18; 95% CI, 1.15-1.22).

**Conclusions:** We created a personalized risk prediction calculator to identify candidates for early vaccine and therapeutics allocation (www.predictcovidrisk.com). These findings may be used to protect those at greatest risk of death from COVID-19.

## Introduction

Early results of new vaccines and therapeutics aimed at fighting the COVID-19 pandemic are promising, however the supply will be constrained given worldwide demand.^1,2^ Initial vaccine administration will be limited to high-risk populations and broader availability will likely be staged over time. On therapeutics, convalescent plasma and Remdesivir have already been supply constrained, placing clinicians in the difficult position of rationing the supply based on risk factors that have not yet been fully elucidated.^3^ This dilemma will be further magnified with the market introduction of monoclonal and polyclonal antibodies which have not been pre-produced as a part of Operation Warp Speed.^4^

Advanced age is a well-known risk factor of COVID-19 death, however, prioritization for vaccine and therapeutic medication administration would be better informed by more complete data.^1,2^ Using a machine learning approach, we explored a wide range of factors associated with COVID-19 death in a cohort of 534,023 COVID-19 patients over 65 years of age. A prediction calculator was created to help identify the personal risk of COVID-19 mortality given an individual’s age, demographic information, and comorbidity profile.

## Methods

### Data source and study population

Through a special data arrangement with the U.S. Centers for Medicare and Medicaid Services (CMS), we accessed the Medicare data server which contains 100% Medicare fee-for-service claims in the U.S. We identified people age 65 years or older who were diagnosed with COVID-19 from carrier, inpatient, and outpatient claims between April 1, 2020 to August 31, 2020. A diagnosis of COVID-19 was identified using *the International Statistical Classification of Disease and Related Health Problem, Tenth Revision* (ICD-10) code U07.1 (2019-nCoV acute respiratory disease).^5^ The Medicare Master Beneficiary Summary File (MBSF), which includes beneficiary enrollment information, was used to identify a patient’s demographic characteristics and geographic location.

### Outcome variables

The primary outcome for this analysis was mortality among hospitalized patients who were diagnosed with COVID-19. Patient death was identified if either of the following two criteria were found in a patient’s inpatient claims: 1) a discharge status was listed as “expired” or 2) a patient status indicator was listed as “died”.

### Independent variables

Patients’ sex, age, race, and geographical location were identified from MBSF. Race was classified as White, Black, Hispanic, Asian, which included Pacific Islander, and Other/Unknown, which included American Indian/Alaska Native, other, unknown. We categorized age groups as 65-69, 70-74, 75-79, 80-84, and 85 years or older. Residential zip codes were used to account for geographic differences.

Comorbidities of patients were captured using the CMS Chronic Conditions Data Warehouse (CCW). The CMS CCW is a database that lists each of the 67 available comorbidity diagnoses that have been assigned to a Medicare beneficiary between January 1, 1999 and December 31, 2018.^6^

### Statistical analysis

We conducted a random forest model, which is a machine learning tool to predict an outcome by creating numerous uncorrelated decision trees to incorporate randomness as a “forest” of trees. During this procedure, the contribution of each predictor towards the predictive accuracy of the resulting model was evaluated; this quantity was referred to as the variable importance. We used the variable importance as an auxiliary means to identify the clinical factors with the highest contributions to the predictive accuracy of the model. Hence, by using a random forest model, we were able to capture conditions and comorbidities that were not emphasized in prior studies. Our adjusted multivariate model included comorbidities identified from the random forest model, comorbidities with a prevalence greater than 30%, and comorbidities that had been reported in previous studies. Geographic clustering was performed by using a logistic multilevel random intercepts model of mortality with patients nested within counties. Mixed-effects multivariable logistic regression with a random intercept for the county was conducted to identify the odds ratio of death. Odds ratios and 95% confidence intervals were reported for each risk factor. A personalized risk prediction nomogram was constructed based on the coefficients from the multivariable logistic model to calculate relative risk using the following formula: probability□=□exp^(_∑_β_×_X)^/[1+ exp^(_∑_β_×_X)^]. The odds ratio was statistically significant at α=0.05 level. Statistical analysis for this study was performed in SAS enterprise version 7.1 (SAS, Inc., Cary, NC).

## Results

### Characteristics of patients diagnosed with COVID-19

From April 1, 2020 through August 20, 2020, 534,023 Medicare beneficiaries over the age of 65 had a diagnosis of 2019 novel coronavirus (COVID-19) in at least one Medicare claim during the study period. Of those these, 148,151 (27.7%) patients were hospitalized and 38,066 (7.1%) died in a hospital.

The highest prevalence of COVID-19 infections was among white patients (n=396,198, 74.2%), female patients (n=307,595, 57.6%), and those age 85 or older (n=138,195, 25.9%). The median age of a COVID-19 diagnosed patient was 77 years (IQR 70-85), and the median age of patients who died was 80 years (IQR 73-87). Each of six comorbidities was present in the majority of COVID-19 patients: 80% (n=423,808) had hypertension, 76% (n=402,979) had hyperlipidemia, 63% (n=335,413) had anemia, 62% (n=332,422) had a cataract, 61% (n=325,498) had rheumatoid arthritis/osteoarthritis, and 54% (n=286,025) had ischemic heart disease (Table 1). Over 65% of patients that died (n=24,927) had at least one of these comorbidities. Risk factors associated with COVID-19 death which had a low prevalence in the Medicare population over 65 years of age included pressure ulcers and chronic ulcers (n=85,740), tobacco use (n=69,437), Schizophrenia and other psychotic disorders (n=65,303), history of acute myocardial infarction (n=29,728), Blindness and Visual Impairment (n=15,202), lymphoma and leukemia (n=11,996), lung cancer (n=7,578), cerebral palsy (n=3,135) sickle cell disease (n=222), For patients over the age of 65 with no comorbidities (n=54,669), the COVID-19 infection fatality rate was 4.7% (n=2,591/54,669).

**Table 1.**
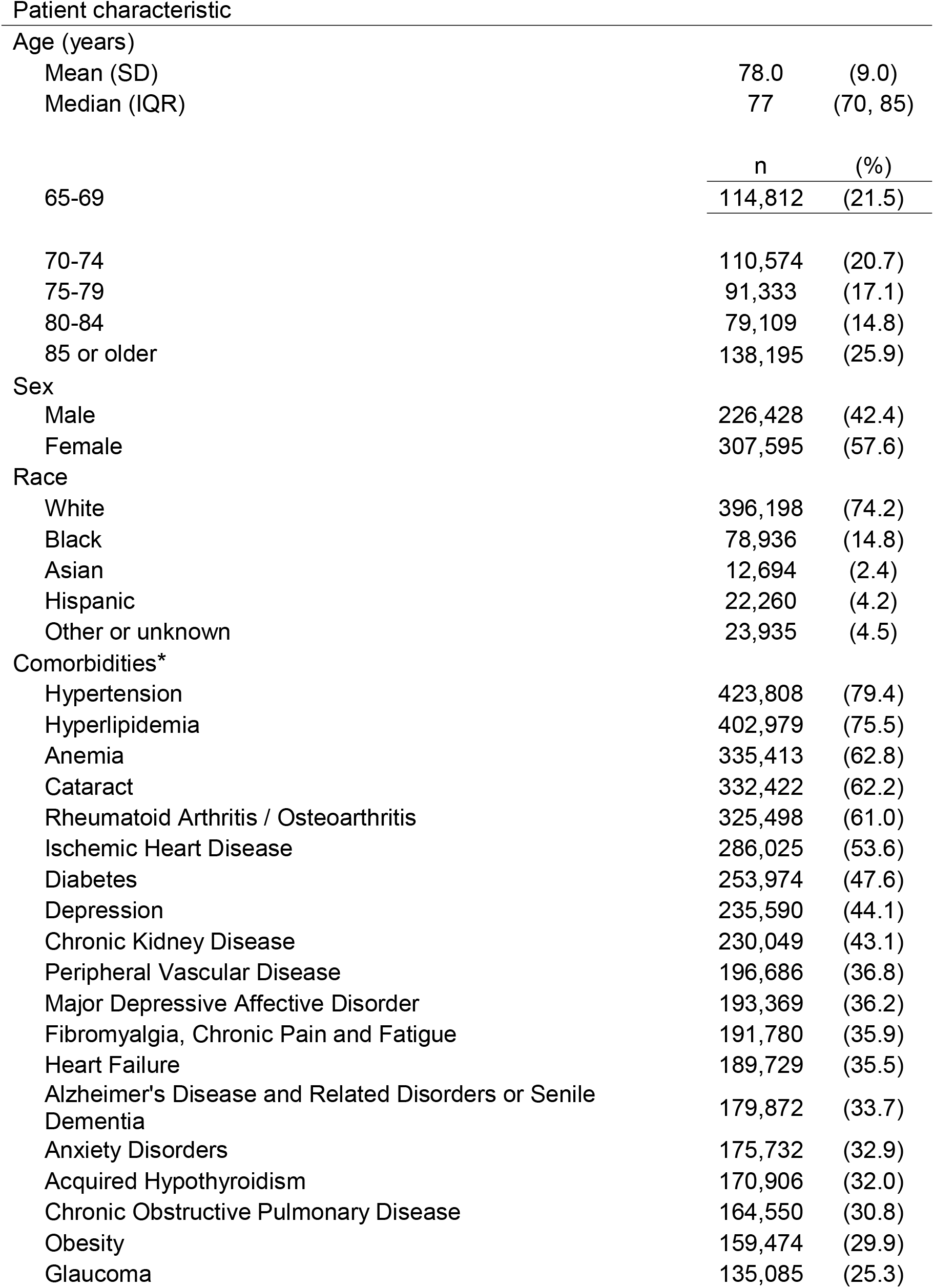
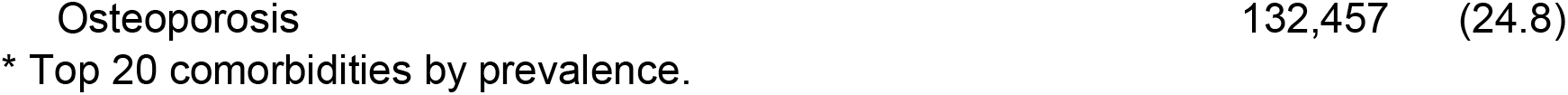
Characteristics of Medicare beneficiaries age 65 years and older diagnosed with COVID-19 from April 1 to August 31, 2020 (n=534,023)

| Patient characteristic |  |  |
| --- | --- | --- |
| Age (years) |  |  |
| Mean (SD) | 78.0 | (9.0) |
| Median (IQR) | 77 | (70, 85) |
|  | n | (%) |
| 65-69 | 114,812 | (21.5) |
| 70-74 | 110,574 | (20.7) |
| 75-79 | 91,333 | (17.1) |
| 80-84 | 79,109 | (14.8) |
| 85 or older | 138,195 | (25.9) |
| Sex |  |  |
| Male | 226,428 | (42.4) |
| Female | 307,595 | (57.6) |
| Race |  |  |
| White | 396,198 | (74.2) |
| Black | 78,936 | (14.8) |
| Asian | 12,694 | (2.4) |
| Hispanic | 22,260 | (4.2) |
| Other or unknown | 23,935 | (4.5) |
| Comorbidities* |  |  |
| Hypertension | 423,808 | (79.4) |
| Hyperlipidemia | 402,979 | (75.5) |
| Anemia | 335,413 | (62.8) |
| Cataract | 332,422 | (62.2) |
| Rheumatoid Arthritis / Osteoarthritis | 325,498 | (61.0) |
| Ischemic Heart Disease | 286,025 | (53.6) |
| Diabetes | 253,974 | (47.6) |
| Depression | 235,590 | (44.1) |
| Chronic Kidney Disease | 230,049 | (43.1) |
| Peripheral Vascular Disease | 196,686 | (36.8) |
| Major Depressive Affective Disorder | 193,369 | (36.2) |
| Fibromyalgia, Chronic Pain and Fatigue | 191,780 | (35.9) |
| Heart Failure | 189,729 | (35.5) |
| Alzheimer's Disease and Related Disorders or Senile Dementia | 179,872 | (33.7) |
| Anxiety Disorders | 175,732 | (32.9) |
| Acquired Hypothyroidism | 170,906 | (32.0) |
| Chronic Obstructive Pulmonary Disease | 164,550 | (30.8) |
| Obesity | 159,474 | (29.9) |
| Glaucoma | 135,085 | (25.3) |

### Variable importance from random forest

A random forest model identified an accurate classification rate of 92.9%, and risk factors that are more likely to be a good fit in a model to predict COVID-19 mortality higher than the norm were chronic kidney disease, prostate cancer, the patient’s race, pressure ulcers and chronic ulcers, acute myocardial infarction, the patient’s sex, and heart failure. We included 20 comorbidities with highest variable importance in the multivariate regression.

### Factors associated with in-hospital death

In an adjusted multivariate model, COVID-19 death was associated with advanced age (85 or older vs 65-69 years, adjusted odds ratio [aOR] = 2.07, CI 1.99-2.16), male sex (aOR = 1.88, CI 1.82-1.94), and non-white race (Hispanic vs White, aOR, =1.74, CI 1.66-1.83; Asian vs White, aOR = 1.71, CI 1.61-1.82; Black vs White, aOR = 1.61, CI 1.56-1.66; Other or unknown vs White, aOR = 1.44, CI 1.37-1.52). Random intercept of the county showed that COVID-19 deaths were clustered by geographic location (p<0.0001). Comorbidities associated with the highest mortality risk included sickle cell disease (aOR = 1.73, CI 1.22-2.47), chronic kidney disease (aOR = 1.32, CI 1.29-1.36), leukemias and lymphomas (aOR =1.22; 95% CI 1.14-1.30), and heart failure (aOR, 1.19, CI 1.16-1.22), followed by diabetes (aOR = 1.18, CI 1.15-1.22) and cerebral palsy (aOR = 1.18, CI 1.04-1.35) (Table 2).

**Table 2.**
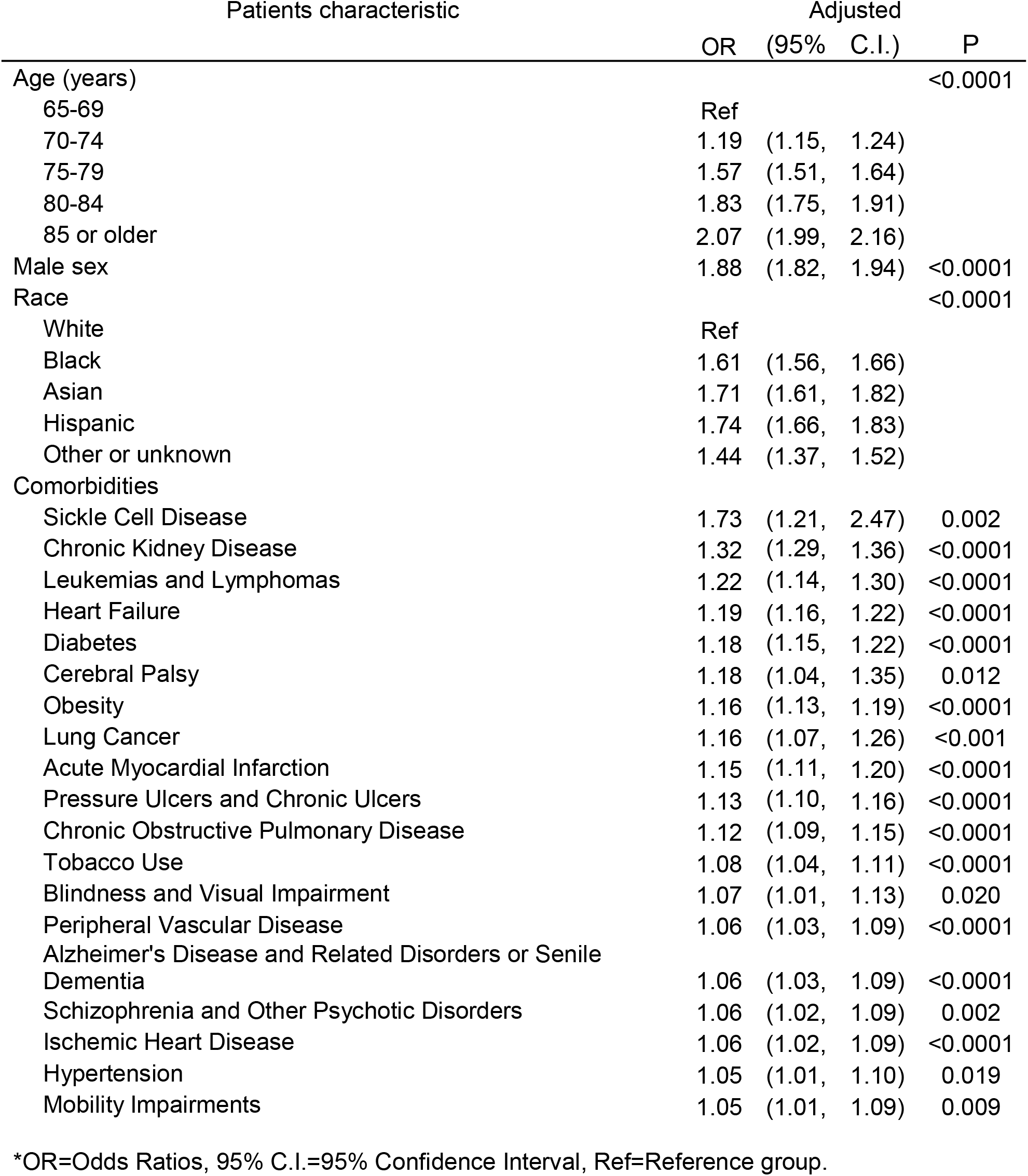
Odds Ratios for COVID-19 death among 534,023 patients^*^.

### Construction of a Risk Calculator

We used the coefficients of patients’ age, sex, race, and all reported risk factors to create a personalized COVID-19 mortality risk prediction calculator. Based on this calculator, an 80-year-old Hispanic man with chronic kidney disease has a mortality risk 6 times higher than a 66-year-old white woman with no comorbidities. The risk calculator is available at www.predictcovidrisk.com.

## Discussion

Our study is the largest comorbidity analysis of COVID-19 patients in the U.S to date. Using a large sample size, geographic clustering, and the encompassing pool of independent variables captured in this study, we identified that the comorbidities of sickle cell disease, chronic kidney disease, leukemias and lymphomas, heart failure, and diabetes are all associated with higher rates of COVID-19 death. These results revealed high risk individuals who should be considered for prioritization of vaccine and therapeutic medication allocation. These findings were also used to develop a risk calculator to allow clinicians to identify patients who were at a highest risk of COVID-19 mortality.

Our results expanded on findings reported in prior studies describing COVID-19 mortality risk factors.^7,8,9^ A large UK-based cohort study of 10,926 COVID-19 deaths found that increased age, male sex, and Black race were associated with a higher risk of mortality.^10^ Our results showed that with every five-year increase over age 65 years, there was an approximately 20% increase in mortality. As consistent with prior reports, we also found racial and ethnic minority groups had a higher risk of COVID-19 mortality compared with White patients. A study in New York of 4,312 COVID-19 patients observed that a Black patients had the highest relative risk compared to other race groups.^11^ Other analyses have found Black patients to have the greater risk of COVID-19 death, however we found that Hispanic patients had the highest mortality risk and that black patients were a close second in this Medicare population.^11-13^ The disproportionate impact of COVID-19 on minorities may be related three possibilities. First, there may be a disproportionate burden of undiagnosed comorbidities such as diabetes, HIV, liver disease, cardiovascular disease, asthma, and kidney disease in minority communities.^14^ Second, minority patients may be acquiring the infection with a higher viral load given denser settings in which minority populations live, commute and work. The size of the viral load involved in a patient has been associated with the infection fatality rate.^15,16^ These populations may also be more likely to live in multigenerational households where ventilation may be poor and effective social distancing may not be feasible.^17^ Finally, minorities may have poorer access to quality health care.^11-13^

Another U.K. study of 20,133 UK patients hospitalized with COVID-19 observed that death was associated with the pre-existing comorbidities of chronic cardiac disease, chronic non-asthmatic pulmonary disease, chronic kidney disease, obesity and liver disease.^18^ Similarly, a U.S. cohort study of 11,721 hospitalized patients with COVID-19 in 38 states found the comorbidities of chronic kidney disease and cardiovascular disease to be associated with an increased odds of mortality.^19^ A U.S.-based study of 521 patients with chronic kidney disease who became critically ill with COVID-19 described a hazard ratio of 1.25.^20^ Our study strongly reinforces the hypothesis that chronic kidney disease is major risk factor for COVID-19 death. We observed that chronic kidney disease was the second leading risk factor among all comorbidities with an adjusted odds ratio of 1.32, second only to sickle cell disease which is rare in the Medicare population over age 65 years.

Using a large sample size, our study affirmed the association of hypertension, diabetes, chronic obstructive pulmonary disease, cardiovascular disease, obesity, and lung cancer with COVID-19 death, in line with previous large size comorbidities studies.^9-11,21-26^ However, different from prior studies, our sample revealed sickle cell disease and leukemias and lymphomas to be major risk factors for COVID-19 death. Previous case studies have reported that sickle cell disease patients had favorable outcomes and suggested that mortality risk in this population was inconclusive.^27,28^ One explanation for our finding may be that sickle cell disease is associated with impaired oxygen exchange, which may be further impeded during the inflammatory phase of the infection.

Obesity has been well-described to be a risk factor for COVID-19 death.^10,18,19^ One study showed that COVID-19 patients with a BMI between 30-34 kg/m2 and >35 kg/m^2^ were 1.8 and 3.6 times more likely to require critical care, respectively, when compared to individuals with a BMI of <30 kg/m^2^.^29^ We observed a risk lower than described in other studies. We believe the weaker association we report to be due to the well-known low sensitivity of obesity codes in claims data to detect obese and overweight individuals.^30^ Therefore, we believe the impact of obesity, overweight, and undocumented metabolic syndrome or pre-diabetes on poor COVID-19 outcomes to be under-reported in this claims based study.

Some of the risk factors we identified may be co-linear with institutionalized patients. Specifically, cerebral palsy, chronic ulcers, blindness, Alzheimer’s disease and related dementias, and mobility impairments are likely more prevalent in persons living with assisted or nursing care. Higher risk in these populations may be due to the mode of transmission in residential facilities.

We reported the first COVID-19 risk factor analysis using machine learning algorithms. Various studies have established the random forest approach as a useful method for modelling risk prediction.^31-33^ We incorporated a random forest approach into the COVID-19 risk factor exploration to identify variables that could be used to predict a better model. Our results revealed that 60% of comorbidities (n=12) captured from random forest model were also in the list of top 20 comorbidities had the highest odds ratio of COVID-19 death. Overall, random forest as a machine learning approach had a preponderance on selecting important variables that could improve the model fitness.^34^

This study had some important limitations. First, we were unable to make conclusions about the infection fatality rate in the community because claims data are specific but not sensitive for infection with COVID-19. Second, we only included inpatient hospital deaths since there is a time lag in the availability of death data outside the hospital setting in the Medicare dataset.

## Conclusion

As COVID-19 vaccines and therapeutics become available, the risk factors we report could inform the allocation of these limited resources until they become more widely available. The application of these results to the personalized risk calculator may help educate clinicians and the public on which patients aged 65 years or older are at highest risk of COVID-19 mortality.

## Data Availability

All data in the manuscript are available from Centers for Medicare & Medicaid Services.

## Acknowledgement

We thank West Health Institute for their research support on this study. We also thank Dr. Brian W. Weir and Dominique Vervoort for their help in preparing this manuscript.

## Author Contributions

Dr. Martin A. Makary had full access to all the data in the study and takes responsibility for the integrity of the data and the accuracy of the data analysis. *Study concept and design*: Makary, Dun, Lash, Segev, Walsh, Bae. *Acquisition, analysis, and interpretation of data*: Makary, Dun, Segev, Walsh, Toner, Bae. *Drafting of the manuscript*: Dun, Makary, Walsh. *Critical revision of the manuscript of important intellectual content*: Makary, Walsh, Segev, Adalja, Toner. Statistical analysis: Dun, Bae. *Obtained funding*: Dun, Walsh, Makary. *Administrative, technical, or material support*: Dun, Walsh, Bae, Adalja, Toner, Lash Hashim, Paturzo, Segev, Makary. *Study supervision*: Makary.

## References

1. Anderson EJ, Rouphael NG, Widge AT, et al. Safety and Immunogenicity of SARS-CoV-2 mRNA-1273 Vaccine in Older Adults. New England Journal of Medicine 2020;0(0):null.

2. Folegatti PM, Ewer KJ, Aley PK, et al. Safety and immunogenicity of the ChAdOx1 nCoV-19 vaccine against SARS-CoV-2: a preliminary report of a phase 1/2, single-blind, randomised controlled trial. The Lancet 2020;396(10249):467–78.

3. Nathan-Kazis J. Gilead’s Remdesivir Supply Will Fall Short of U.S. Need This Summer, Analyst Says [Internet]. [cited 2020 Oct 22];Available from: https://www.barrons.com/articles/gilead-remdesivir-supply-donate-covid-19-treatment-emergency-use-fda-51589202131

4. US Department of Health and Human Services (2020). Explaining Operation Warp Speed. US Department of Health and Human Services. [cited 2020 Oct 22];Available from https://www.hhs.gov/sites/default/files/fact-sheet-operation-warp-speed.pdf

5. WHO | Emergency use ICD codes for COVID-19 disease outbreak [Internet]. WHO. [cited 2020 Oct 22];Available from: http://www.who.int/classifications/icd/covid19/en/

6. Condition Categories - Chronic Conditions Data Warehouse [Internet]. [cited 2020 Oct 22];Available from: https://www2.ccwdata.org/web/guest/condition-categories

7. Zhou F, Yu T, Du R, et al. Clinical course and risk factors for mortality of adult inpatients with COVID-19 in Wuhan, China: a retrospective cohort study. The Lancet 2020;395(10229):1054–62.

8. Lipsitch M, Swerdlow DL, Finelli L. Defining the Epidemiology of Covid-19 — Studies Needed. New England Journal of Medicine 2020;382(13):1194–6.

9. Imam Z, Odish F, Gill I, et al. Older age and comorbidity are independent mortality predictors in a large cohort of 1305 COVID-19 patients in Michigan, United States. Journal of Internal Medicine 2020;288(4):469–76.

10. Williamson EJ, Walker AJ, Bhaskaran K, et al. Factors associated with COVID-19-related death using OpenSAFELY. Nature 2020;584(7821):430–6.

11. Golestaneh L, Neugarten J, Fisher M, et al. The association of race and COVID-19 mortality. EClinicalMedicine [Internet] 2020 [cited 2020 Oct 22];25. Available from: https://www.thelancet.com/journals/eclinm/article/PIIS2589-5370(20)30199-1/abstract

12. Price-Haywood EG, Burton J, Fort D, Seoane L. Hospitalization and Mortality among Black Patients and White Patients with Covid-19. New England Journal of Medicine 2020;382(26):2534–43.

13. Tai DBG, Shah A, Doubeni CA, Sia IG, Wieland ML. The Disproportionate Impact of COVID-19 on Racial and Ethnic Minorities in the United States. Clin Infect Dis [Internet] [cited 2020 Oct 22];Available from: https://academic.oup.com/cid/advance-article/doi/10.1093/cid/ciaa815/5860249

14. Webb Hooper M, Nápoles AM, Pérez-Stable EJ. COVID-19 and Racial/Ethnic Disparities. JAMA 2020;323(24):2466.

15. Declining Trend in the Initial SARS-CoV-2 Viral Load final.mp4 [Internet]. [cited 2020 Oct 22]. Available from: https://drive.google.com/file/d/10AWAhVurFq2-R8AIvjC3sEMaN58xiLrG/view?usp=embed_facebook

16. Piubelli C, Deiana M, Pomari E, et al. Overall decrease of SARS-CoV-2 viral load and reduction of clinical burden: the experience of a Northern Italy hospital. Clinical Microbiology and Infection [Internet] 2020 [cited 2020 Oct 22];0(0). Available from: https://www.clinicalmicrobiologyandinfection.com/article/S1198-743X(20)30617-0/abstract

17. CDC. Communities, Schools, Workplaces, & Events [Internet]. Centers for Disease Control and Prevention. 2020 [cited 2020 Oct 22];Available from: https://www.cdc.gov/coronavirus/2019-ncov/community/index.html

18. Docherty AB, Harrison EM, Green CA, et al. Features of 20 133 UK patients in hospital with covid-19 using the ISARIC WHO Clinical Characterisation Protocol: prospective observational cohort study. BMJ [Internet] 2020 [cited 2020 Oct 22];369. Available from: https://www.bmj.com/content/369/bmj.m1985

19. Fried MW, Crawford JM, Mospan AR, et al. Patient Characteristics and Outcomes of 11 721 Patients With Coronavirus Disease 2019 (COVID-19) Hospitalized Across the United States. Clin Infect Dis [Internet] [cited 2020 Oct 22];Available from: https://academic.oup.com/cid/advance-article/doi/10.1093/cid/ciaa1268/5898276

20. Flythe JE, Assimon MM, Tugman MJ, et al. Characteristics and Outcomes of Individuals With Pre-existing Kidney Disease and COVID-19 Admitted to Intensive Care Units in the United States. Am J Kidney Dis [Internet] 2020 [cited 2020 Oct 22];Available from: https://www.ncbi.nlm.nih.gov/pmc/articles/PMC7501875/.

21. Guan W, Liang W, Zhao Y, et al. Comorbidity and its impact on 1590 patients with Covid-19 in China: A Nationwide Analysis. European Respiratory Journal [Internet] 2020 [cited 2020 Oct 22];Available from: https://erj.ersjournals.com/content/early/2020/03/17/13993003.00547-2020

22. Yang J, Zheng Y, Gou X, et al. Prevalence of comorbidities and its effects in patients infected with SARS-CoV-2: a systematic review and meta-analysis. Int J Infect Dis 2020;94:91–5.

23. Noor FM, Islam MdM. Prevalence and Associated Risk Factors of Mortality Among COVID-19 Patients: A Meta-Analysis. J Community Health 2020;1–13.

24. Fang X, Li S, Yu H, et al. Epidemiological, comorbidity factors with severity and prognosis of COVID-19: a systematic review and meta-analysis. Aging (Albany NY) 2020;12(13):12493–503.

25. Zheng Z, Peng F, Xu B, et al. Risk factors of critical & mortal COVID-19 cases: A systematic literature review and meta-analysis. Journal of Infection 2020;81(2):e16–25.

26. Maciel EL, Jabor P, Goncalves Júnior E, et al. Factors associated with COVID-19 hospital deaths in Espírito Santo, Brazil, 2020. Epidemiol Serv Saude 2020;29(4):e2020413.

27. Kehinde TA, Osundiji MA. Sickle cell trait and the potential risk of severe coronavirus disease 2019—A mini-review. Eur J Haematol [Internet] 2020 [cited 2020 Oct 22];Available from: https://www.ncbi.nlm.nih.gov/pmc/articles/PMC7361772/

28. McCloskey KA, Meenan J, Hall R, Tsitsikas DA. COVID-19 infection and sickle cell disease: a UK centre experience. British Journal of Haematology 2020;190(2):e57–8.

29. Lighter J, Phillips M, Hochman S, et al. Obesity in Patients Younger Than 60 Years Is a Risk Factor for COVID-19 Hospital Admission. Clin Infect Dis 2020;71(15):896–7.

30. Martin B-J, Chen G, Graham M, Quan H. Coding of obesity in administrative hospital discharge abstract data: accuracy and impact for future research studies. BMC Health Serv Res 2014;14:70.

31. Yeşilkanat CM. Spatio-temporal estimation of the daily cases of COVID-19 in worldwide using random forest machine learning algorithm. Chaos Solitons Fractals 2020;140:110210.

32. Cutler DR, Edwards TC, Beard KH, et al. Random Forests for Classification in Ecology. Ecology 2007;88(11):2783–92.

33. Jeung M, Baek S, Beom J, Cho KH, Her Y, Yoon K. Evaluation of random forest and regression tree methods for estimation of mass first flush ratio in urban catchments. Journal of Hydrology 2019;575:1099–110.

34. Weng SF, Reps J, Kai J, Garibaldi JM, Qureshi N. Can machine-learning improve cardiovascular risk prediction using routine clinical data? PLOS ONE 2017;12(4):e0174944.

